# Self-collection and pooling of samples as resources-saving strategies for RT-PCR-based SARS-CoV-2 surveillance, the example of travelers in French Polynesia

**DOI:** 10.1101/2021.06.17.21254195

**Authors:** Maite Aubry, Iotefa Teiti, Anita Teissier, Vaea Richard, Teheipuaura Mariteragi-Helle, Kiyojiken Chung, Farah Deen, Tuterarii Paoaafaite, Van-Mai Cao-Lormeau

**Author notes:** **Corresponding author:** (MA).

## Abstract

In French Polynesia, the first case of SARS-CoV-2 infection was detected on March 10^th^, 2020, in a resident returning from France. Between March 28^th^ and July 14^th^, international air traffic was interrupted and local transmission of SARS-CoV-2 was brought under control, with only 62 cases recorded. The main challenge for reopening the air border without requiring travelers to quarantine on arrival was to limit the risk of re-introducing SARS-CoV-2. Specific measures were implemented, including the obligation for all travelers to have a negative RT-PCR test for SARS-CoV-2 carried out within 3 days before departure, and to perform another RT-PCR testing 4 days after arrival. Because of limitation in available medical staff, travelers were provided a kit allowing self-collection of oral and nasal swabs. In addition to increase our testing capacity, self-collected samples from up to 10 travelers were pooled before RNA extraction and RT-PCR testing. When a pool tested positive, RNA extraction and RT-PCR were performed on each individual sample. We report here the results of COVID-19 surveillance (COV-CHECK PORINETIA) conducted between July 15^th^, 2020, and February 15^th^, 2021, in travelers using self-collection and pooling approaches. We tested 5,982 pools comprising 59,490 individual samples, and detected 273 (0.46%) travelers positive for SARS-CoV-2. A mean difference of 1.17 Ct (CI 95% 0.93 – 1.41) was found between positive individual samples and pools (N=50), probably related to the volume of samples used for RNA extraction (200 µL versus 50 µL, respectively). Retrospective testing of positive samples self-collected from October 20^th^, 2020, using variants-specific amplification kit and spike gene sequencing, found at least 6 residents infected by the B1.1.7 UK variant. Self-collection and pooling approaches allowed large-scale screening for SARS-CoV-2 using less human, material and financial resources. Moreover, this strategy allowed detecting the introduction of SARS-CoV-2 variants in French Polynesia.

## Introduction

Cases of respiratory infection (coronavirus disease 2019, COVID-19) caused by the severe acute respiratory syndrome coronavirus 2 (SARS-CoV-2) were first reported in December 2019 in Wuhan City, Hubei Province, China [1]. Because of the disease global expansion, COVID-19 was declared by the World Health Organization as a public health emergency of international concern on January 30^th^, 2020 [2], and was then characterized as a pandemic on March 11^th^ [3].

French Polynesia is a French overseas collectivity in the South Pacific with ca 190,000 inhabitants living on the island of Tahiti, and 90,000 inhabitants distributed on 73 other islands scattered on a surface area as wide as Europe [4]. The first case of SARS-CoV-2 infection was detected on March 10^th^, 2020, in a resident of Tahiti returning from France [5]. To stop viral transmission, the population of French Polynesia was confined and international air traffic was interrupted on March 20^th^ and 28^th^, respectively. Only residents or foreigners showing a compelling reason were allowed to enter French Polynesia provided they were quarantined on arrival. Confinement was eased from April 20^th^ in most islands as no active circulation of SARS-CoV-2 had been detected, then was fully leveraged on May 21^st^. Between March 10^th^ and June 25^th^, among 5,390 patients tested for a suspicion of SARS-CoV-2 infection, a total of 62 were found positive, including 32 imported cases, and no COVID-19 related death was recorded [6].

Tourism is an important financial resource for French Polynesia [7]. Consequently it was urgent to reopen the international air traffic, but without imposing a quarantine on arrival so as not to deter tourists from traveling. At the same time, a strategy needed to be found to protect the population from a new introduction of the virus, especially in remote touristic islands where health facilities are limited. Screening for SARS-CoV-2 all travelers arriving in French Polynesia was impossible because of the lack of medical staff to collect naso-pharyngeal swabs, and of laboratory staff trained in molecular biology to analyze the collected samples. Moreover, the number of available test kits for RNA extraction and RT-PCR assays was limited due to the global shortage.

A prospective study conducted in Australia on 236 ambulatory patients showed that the performance of self-collected throat and nasal swabs was at least equivalent to that of throat and nasal/naso-pharyngeal swabs collected by health workers for the detection of SARS-CoV-2 [8]. Thus, self-collection would offer a reliable alternative to health worker collected samples, and would reduce the need for trained medical staff. Moreover, several studies demonstrated that pooling of samples prior to RNA extraction and RT-PCR testing was reliable compared to the analysis of individual samples for SARS-CoV-2 detection [9-13]. This method proved efficient to increase testing capacity by saving reagents and laboratory-staff worktime, thus allowing large-scale screening of asymptomatic populations.

We describe here the use of self-collected and pooled samples as resources-saving strategies for RT-PCR-based surveillance of SARS-CoV-2 in travelers entering French Polynesia. From July 15^th^, 2020, to February 9^th^, 2021, all travelers had to comply with a testing protocol (COV-CHECK PORINETIA) which combined 2 consecutive RT-PCR tests for the detection of SARS-CoV-2 [14]. The first one was performed within 3 days before departure from an individual oro-and/or naso-pharyngeal sample collected by a health worker. A negative RT-PCR result was required to allow departure. The second RT-PCR test was performed 4 days after arrival from a pool including up to 10 oral and nasal samples self-collected by the travelers. In case of positive RT-PCR, all samples included in the pool were re-tested individually. Moreover, from February 2021, due to the worldwide emergence of SARS-CoV-2 variants of public health concern [15], self-collected samples found positive by SARS-CoV-2 RT-PCR were re-tested using variant-specific amplification kit. Both retrospective and prospective investigations conducted from samples self-collected by travelers over the past four months showed the introduction of the B1.1.7 United Kingdom (UK) variant in French Polynesia.

## Methodology

### Procedure before departure

Within 3 days before departure, each traveler (including minors) had to register online on the « Electronic Travel Information System » platform (ETIS, https://www.etis.pf/), implemented by the government of French Polynesia since July 11^th^, 2020. The ETIS form contained information about the status of the traveler (resident or non-resident of French Polynesia), and personal information including name, gender, date of birth, passport number, mobile phone number (local or international), e-mail address, geographical address in French Polynesia for residents, or country of residence and contact-person in case of emergency for non-residents. The traveler had to declare to have tested negative for SARS-CoV-2 by RT-PCR within 3 days prior to departure, certify to present no symptoms of COVID-19 at boarding, agree to comply with all sanitary rules required by the government of French Polynesia (including wearing a mask in public areas, avoiding close contact with people as much as possible, reporting any symptom suggestive of COVID-19), certify to have a travel insurance for non-French citizens and agree to assume all health costs incurred in French Polynesia, and accept to perform a self-test for SARS-CoV-2 detection using a kit provided upon arrival. Finally, non-resident travelers had to accurately describe their itinerary by indicating arrival and departure dates and flight numbers, and each visited island with the dates of stay as well as the name and contact of the accommodation(s). Data collected on the ETIS platform were stored in accordance with applicable General Data Protection Regulation (GDPR) laws. They could be consulted only by the French Polynesia health authorities and used for the protection of public health and epidemiological research. Once finalized, the traveler received the receipt of the ETIS form at the email address indicated for the registration. Each traveler was identified by a unique ETIS number and a QR code. On the day of departure, the traveler had to present the ETIS receipt and the proof of negative RT-PCR test at check-in to be allowed to board the flight.

### Self-sampling kit distribution on arrival

On arrival at Tahiti-Faa’a international airport (Tahiti), each traveler aged at least 6 years and staying more than 4 days in French Polynesia received a self-sampling kit consisting of a zipper plastic bag containing 2 swabs (one for the oral sampling and the other for the nasal sampling), a tube with 2-3 ml of viral transport medium (VTM) or universal transport medium (UTM), and a plastic bag approved for the transport of bio-hazardous materials with an absorbent inside. With the kit were also provided in both French and English languages an information notice presenting the protocol implemented for COVID-19 surveillance of travelers, an instruction sheet describing how to proceed to sample self-collection, and the list of the health care centers located on the different islands of French Polynesia where self-collected samples could be dropped off (S1 File).

The date of completion of the self-collection of samples was indicated on the kit and corresponded to the 4^th^ day after arrival in French Polynesia, excluding weekends because of the closure of most health care centers (if the 4^th^ day fell on a Saturday, the test was brought forward to the previous Friday, and if it fell on a Sunday the test was postponed to the following Monday). The choice of the 4^th^ day for sample self-collection was based on the mean incubation period, *ie* the delay between the exposure to the virus and the apparition of symptoms of COVID-19, which had been estimated to 5-6 days in previously published studies [16-18]. Since the first RT-PCR testing had to be performed within 3 days prior to departure, the delay between possible infection and detectable viremia would correspond to 2-6 days (mean 4 days) after arrival in French Polynesia.

Each kit was identified by a unique number represented by a barcode affixed to both the zipper plastic bag and the tube containing the transport medium. Prior to giving the kit to the traveler, the kit number was matched to the traveler’s ETIS number using an application installed on mobile digital tools (tablet or smartphone) that successively scanned the barcode on the kit and the QR code on the traveler’s ETIS receipt. Those data were sent automatically to the server of the ETIS platform to be associated with the traveler’s identity. The data allowing the link between the kit number and the identity of the traveler could only be viewed by the physicians of the Surveillance office from the Ministry of Health of French Polynesia.

### Sample self-collection and shipment

Two days after arrival, each traveler received an automatic message from the tourism department of French Polynesia at the e-mail address provided in the ETIS form, as a reminder to perform samples self-collection on the date indicated on the kit. Travelers could refer to the instruction sheet received upon arrival and to the video tutorial posted online by the tourism department (available at: https://www.youtube.com/watch?v=pYcOp6tk9bo&feature=emb_logo). Moreover, a call center open 7 days a week and an e-mail address were available to answer travelers’ questions.

In order to increase the probability of collection of viral particles at the respiratory tract, the traveler had to collect both nasal and oral samples. Briefly, the traveler had to insert the swab provided for the nasal sampling into each nostril until some resistance was felt, and rub the walls 4 times (the swab provided for the nasal sampling had a large cotton tip to prevent the traveler from pushing it too deep and getting hurt). In addition, the traveler had to insert the swab provided for the oral sampling into the mouth and rub several times inside of cheeks, the top and the bottom of the tongue, the palate, and the lower and upper gums. Then, the traveler had to insert the 2 swabs into the tube containing the transport medium and close it, turn the tube 3 times to mix the samples with the medium, put the tube into the bio-hazard plastic bag and seal it, and finally put the bio-hazard plastic bag back into the zipper plastic bag.

The traveler had to keep the self-collected samples at a temperature between +4°C and +8°C until dropping it off at the Institut Louis Malardé (ILM, Papeete, Tahiti) or one of the health care centers on the list provided with the kit. Travelers staying in a hotel, guesthouse or on a cruise ship participating in the surveillance strategy also had the opportunity to drop off their self-collected samples at the lobby of their accommodation. Then, the accommodation was responsible for forwarding the self-collected samples to the nearest health care center under the temperature conditions recommended during transport. In Tahiti, self-collected samples were collected from health care centers every weekday. In the other islands, self-collected samples dropped off at the health care centers were shipped by boat or plane to Tahiti once to several times a week, depending on the frequency of sea and air rotations (Fig 1). All self-collected samples were ultimately delivered to ILM.

**Fig 1:**
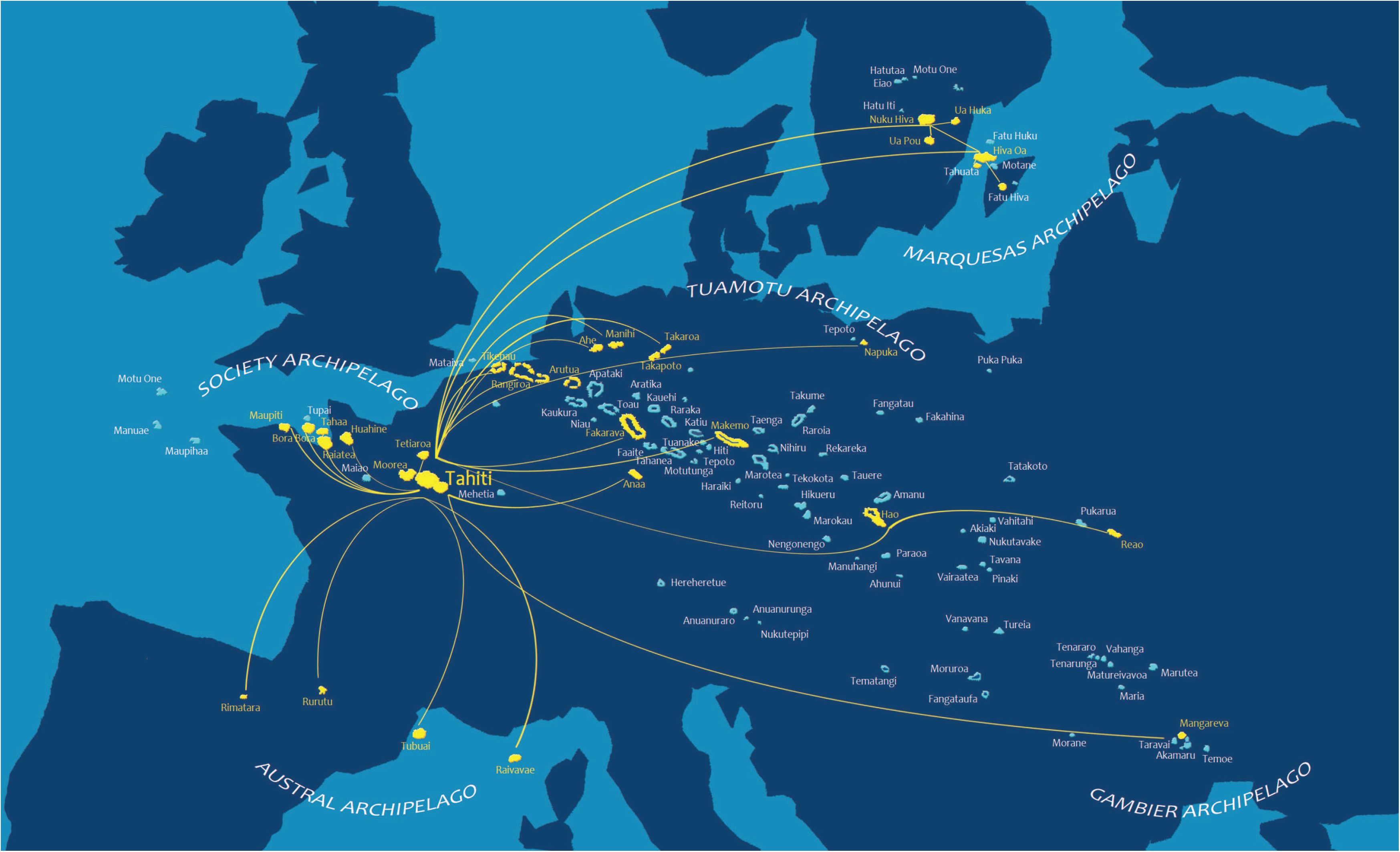
Air and maritime connections for the shipment of travelers’ self-collected samples from the islands with health care centers participating in the COV-CHECK PORINETIA surveillance strategy, to Tahiti where the Institut Louis Malardé is located; a surface area as wide as Europe.

### Analysis of self-collected samples

#### Pool size validation

In order to increase testing capacity for SARS-CoV-2 detection at ILM, we assessed the use of pooled samples from different travelers before viral RNA extraction and RT-PCR steps. A previous study had shown that 8 to 32 individual samples could be pooled without compromising the benefit of the pooling strategy for a SARS-CoV-2 prevalence ranging from 2 to 0.1 % [19]. Since travelers entering French Polynesia had a negative SARS-CoV-2 RT-PCR test dating less than 3 days before departure, the expected prevalence among this population was low.

In order to assess the use of a pool size of 10 individual samples, we performed 10-fold serial dilutions of a nasopharyngeal sample collected from a patient tested RT-PCR positive for SARS-CoV-2 in French Polynesia in March 2020 (Fig 2). Then, we mixed 50 µL of each 10-fold dilution with 50 µL of 9 nasopharyngeal samples collected from patients tested RT-PCR negative for SARS-CoV-2, with a final volume of 500 µL per pool. We subsequently performed viral RNA extraction and RT-PCR.

**Fig 2:**
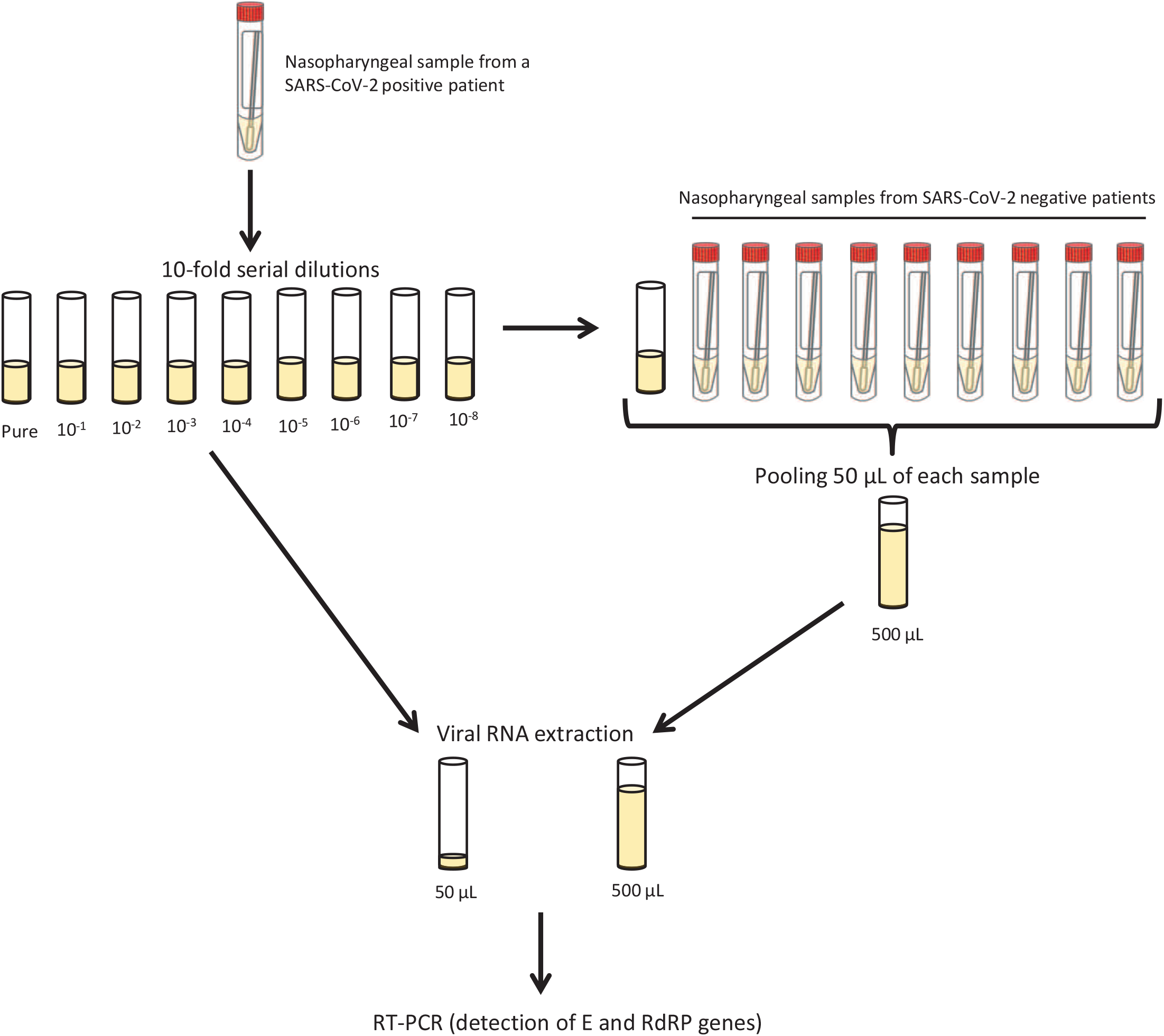
Protocol for the evaluation of the pooling strategy.

Viral RNA extraction was performed on the NucliSENS® easyMag® instrument (bioMérieux, France). A volume of 50 µL of each 10-fold dilution or 500 µL of each pool was mixed with 50 µL or 100 µL of magnetic silica, respectively. The final elution volume was set up at 50 µL. To check the validity of the extraction process, 10 µL of internal positive extraction control (LightMix Modular EAV RNA Extraction Control kit, TIB MOLBIOL, Germany) were added to each dilution or pool.

Duplex RT-PCR was performed on the CFX96 Touch Real-Time PCR Detection System (Bio-Rad, USA), using 2 different protocols.

In the first protocol, 7.5 µL of each extracted RNA were added to 12.5 µL of reaction mixture containing 10 µL of 2X Reaction buffer, 0.32 µL of MgSO4, and 0.8 µL of RT/Taq mixture provided with SuperScript™ III Platinum™ One-Step qRT-PCR Kit (Invitrogen, USA), 0.5 µL of primers and probe mixture from LightMix® Modular SARS and Wuhan CoV E gene kit or from LightMix® Modular Wuhan CoV RdRP-gene kit, and 0.5 µL of primers and probe mixture from LightMix® Modular EAV RNA Extraction Control kit (TIB MOLBIOL, Germany) [19]. Reverse transcription was carried out at 55 °C for 5 min, followed by the initial denaturation and polymerase activation steps at 95 °C for 5 min, 45 cycles of amplification at 95 °C for 5 sec, 60 °C for 15 sec and 72 °C for 5 sec, and a final cooling step at 40 °C for 30 sec.

In the second protocol, 5 µL of each extracted RNA were added to 15 µL of reaction mixture containing 10 µL of 2X Reaction buffer and 0.5 µL of Reverse Transcriptase provided with iTaq Universal Probes One-Step kit (Bio-Rad, USA), 0.5 µL of primers and probe mixture from LightMix® Modular SARS and Wuhan CoV E gene kit or from LightMix® Modular Wuhan CoV RdRP-gene kit, and 0.5 µL of primers and probe mixture from LightMix® Modular EAV RNA Extraction Control kit. Reverse transcription was carried out at 50 °C for 10 min, followed by the initial denaturation and polymerase activation steps at 95 °C for 3 min, and then 45 cycles of amplification at 95 °C for 15 sec and 60 °C for 30 sec.

#### Pooling of samples

Upon receipt at ILM, the barcode of each self-collected sample was scanned, and up to 10 self-collected samples were randomly grouped. A new barcode assigned to the group of self-collected samples was scanned and stuck on a tube containing 2 mL of NucliSENS lysis buffer (BioMérieux, France). Under a class II biological safety cabinet, the tubes containing the transport medium and the 2 swabs used for nasal and oral samples self-collection were removed from the bio-hazard and zipper plastic bags, and vortexed for 30 sec. Then, 50 μL of transport medium were taken from each of the grouped tubes and transferred into the pooling tube containing the lysis buffer. The pooling tube was subsequently vortexed for 10 sec and incubated at room temperature for at least 10 min before the extraction step.

#### RNA extraction and RT-PCR

Viral RNA extraction and duplex RT-PCR were performed from pooled self-collected samples as described above, using LightMix® Modular SARS and Wuhan CoV E gene kit and LightMix® Modular EAV RNA Extraction Control kit. If a pool tested positive, 200 µL of each self-collected sample were individually subjected to viral RNA extraction, by adding 50 µL of magnetic silica and 10 µL of internal positive extraction control. The presence of SARS-CoV-2 RNA was confirmed by the detection of specific RdRP gene using LightMix® Modular Wuhan CoV RdRP-gene kit, with similar conditions of RT-PCR as described above.

### Confirmatory test

Barcode numbers corresponding to individual self-collected samples tested positive were sent to the Surveillance office from the Ministry of Health. Using the data provided on the ETIS platform, travelers were contacted and immediately isolated. A naso-pharyngeal swab was collected from each traveler by a health worker and tested by RT-PCR to confirm SARS-CoV-2 infection.

#### Detection of variants

Individual self-collected samples found positive for SARS-CoV-2 between October, 2020, and February, 2021, were re-tested using the VirSNiP SARS-CoV-2 Spike N501Y Kit (TIB MOLBIOL, Germany) that specifically detects the mutation N501Y found in the spike (S) protein of the B.1.1.7 (UK), B.1.351 (South Africa, SA) and P.1 (Brazil) variants [15], and reagents from SuperScript™ III Platinum™ One-Step qRT-PCR Kit. Briefly, 7.5 µL of extracted RNA were added to 12.5 µL of reaction mixture containing 10 µL of 2X Reaction buffer, 0.30 µL of MgSO4, and 0.8 µL of RT/Taq mixture provided with SuperScript™ III Platinum™ One-Step qRT-PCR Kit, and 0.5 µL of primers and probe mixture from VirSNiP SARS-CoV-2 Spike N501Y Kit. Reverse transcription was carried out at 55 °C for 5 min, followed by the initial denaturation and polymerase activation steps at 95 °C for 5 min, and then 45 cycles of amplification at 95 °C for 5 sec, 60 °C for 15 sec and 72 °C for 5 sec. The presence of the 501Y mutation was revealed by the amplification curve with a melting temperature of 61.2±2 °C.

To identify the variant lineage (UK, SA or Brazil), the complete S gene (nucleotide positions 21,563 to 25,384) was sequenced with the 3500 series genetic analyzer (Applied Biosystems, USA), using primers provided by the Laboratory for Urgent Response to Biological Threats (CIBU) at the Institut Pasteur (Paris, France), and reagents from the Big Dye Terminator V3.1 kit (Applied Biosystems, USA). Partial sequences were cleaned and assembled using the Sequencher 4.10 software (Gene Codes Corporation, USA). The final sequence was uploaded on the GISAID CoVsurver (available at https://www.gisaid.org/epiflu-applications/covsurver-mutations-app/) that automatically determines nucleotide/amino acid mutations and deletions compared to the reference sequence hCoV-19/Wuhan/WIV04/2019.

### Statistical analysis

Data were analyzed with GraphPad Prism 7.04. The paired t test was used to compare RT-PCR results between individual and pooled samples. P values < 0.05 were considered to be significant.

### Ethics

The COV-CHECK PORINETIA surveillance strategy was implemented by the government of French Polynesia (order No. 525 CM of May 13^th^, 2020, amended by order No. 961 CM of July 8^th^, 2020 ; published in full in the Official Journal 2020 No. 79 NS of July 9^th^, 2020). The approval for the use of data produced within the framework of the COV-CHECK PORINETIA surveillance strategy was obtained from the ethics committee of French Polynesia (No. 90 CEPF of June 15^th^, 2021)

## Results

### Validation of the pooling strategy

To assess the impact of pooling samples on SARS-CoV-2 detection sensitivity, we compared RT-PCR results (Ct values) obtained following viral RNA extraction from 50 µL of 10-fold serial dilutions of a positive nasopharyngeal sample, and from 500 µL of pools containing 50 µL of each diluted sample with 50 µL of 9 negative nasopharyngeal samples. Because of the limited number of positive nasopharyngeal samples available at the time the assessment was performed and the short delay before reopening the air border, only one sample could be tested.

Reagents from 2 different RT-PCR kits (SuperScript™ III Platinum™ One-Step qRT-PCR Kit or iTaq Universal Probes One-Step kit) were used to detect the SARS-CoV-2 E and RdRP genes. Whatever the RT-PCR kit used and the gene detected, Ct values were not significantly different between individual and pooled samples, with p-values ranging from 0.051 to 0.699 (Table 1).

**Table 1:** Comparison of individual test Ct value with pool Ct value using 10-fold serial dilutions of a positive nasopharyngeal sample, and 2 different RT-PCR kits for the detection of SARS-CoV-2 E and RdRP genes.

| Dilution | SuperScript™ III Platinum™ One-Step qRT-PCR Kit |  |  |  | iTaQ Universal Probes One-Step kit |  |  |  |
| --- | --- | --- | --- | --- | --- | --- | --- | --- |
|  | E gene detection |  | RdRP gene detection |  | E gene detection |  | RdRP gene detection |  |
|  | Individual | Pool | Individual | Pool | Individual | Pool | Individual | Pool |
| – | 13,67 | 14 | 13,42 | 13,53 | 13,67 | 13,92 | 13,49 | 13,74 |
| 10<sup>-1</sup> | 17,04 | 17,1 | 16,61 | 16,73 | 16,9 | 17,12 | 16,84 | 16,99 |
| 10<sup>-2</sup> | 20,43 | 20,34 | 20,5 | 20,26 | 20,41 | 20,29 | 20,79 | 20,38 |
| 10<sup>-3</sup> | 23,54 | 23,48 | 23,37 | 23,24 | 23,55 | 23,72 | 23,48 | 23,49 |
| 10<sup>-4</sup> | 27,12 | 27,29 | 27,04 | 27,02 | 27,17 | 27,33 | 27,1 | 27,28 |
| 10<sup>-5</sup> | 30,5 | 31,26 | 30,35 | 30,32 | 30,63 | 30,78 | 30,33 | 30,61 |
| 10<sup>-6</sup> | 34,05 | 34,57 | 35,71 | 34,47 | 35,71 | 34,2 | 34,63 | 35,04 |
| 10<sup>-7</sup> | 38,73 | 39,82 | 44,11 | ND | 41,71 | ND | 43,59 | 44,6 |
| 10<sup>-8</sup> | ND | ND | ND | ND | ND | ND | ND | ND |

ND: SARS-CoV-2 RNA not detected.

### Detection of SARS-CoV-2 infection in travelers

From July 15^th^, 2020, to February 15^th^, 2021, we tested 5,982 pools comprising 59,490 individual self-collected samples, and found 238 pools (3.98%) positive for SARS-CoV-2 E gene (S1 Table). RNA extraction and RT-PCR subsequently performed on individual samples confirmed that 273 travelers were positive for SARS-CoV-2 specific RdRP gene detection.

Among the pools tested, 210, 23, 4 and 1 contained respectively 1, 2, 3 and 4 positive individual samples (Figs 3a-c). The prevalence of positive self-collected samples among all travelers tested was 0.46 %.

**Fig 3.**
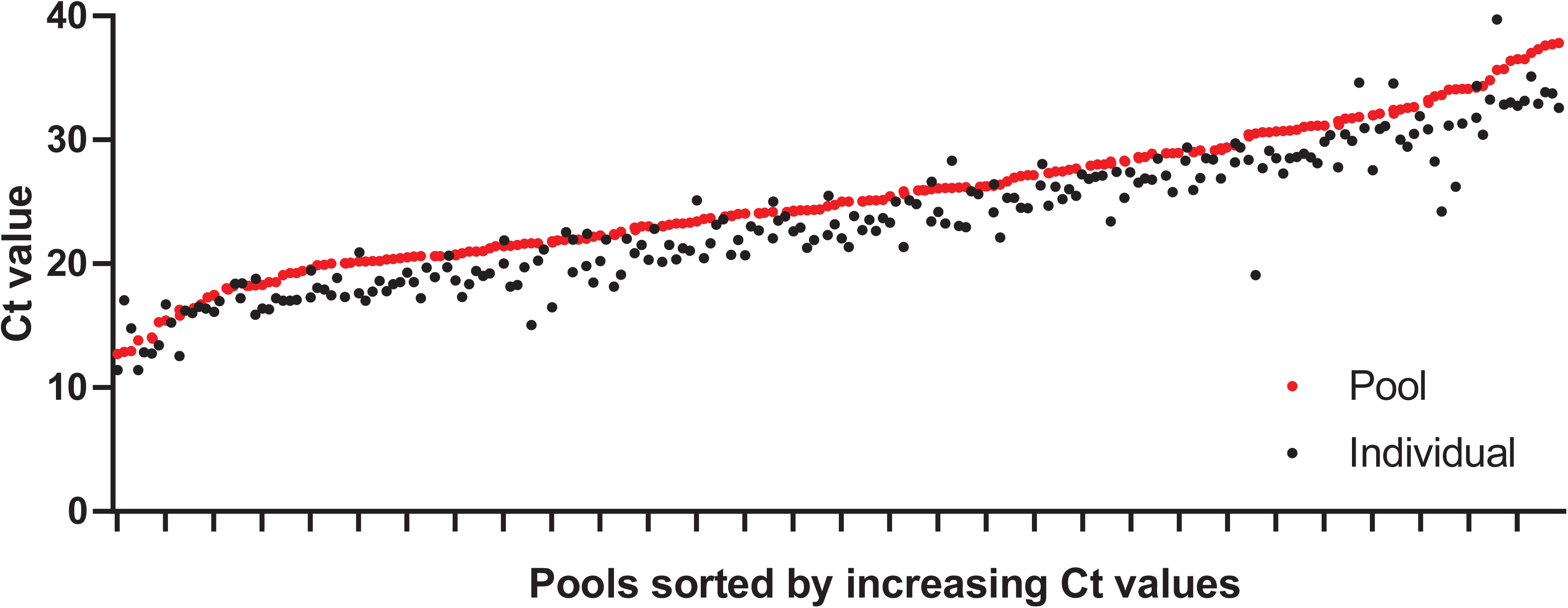
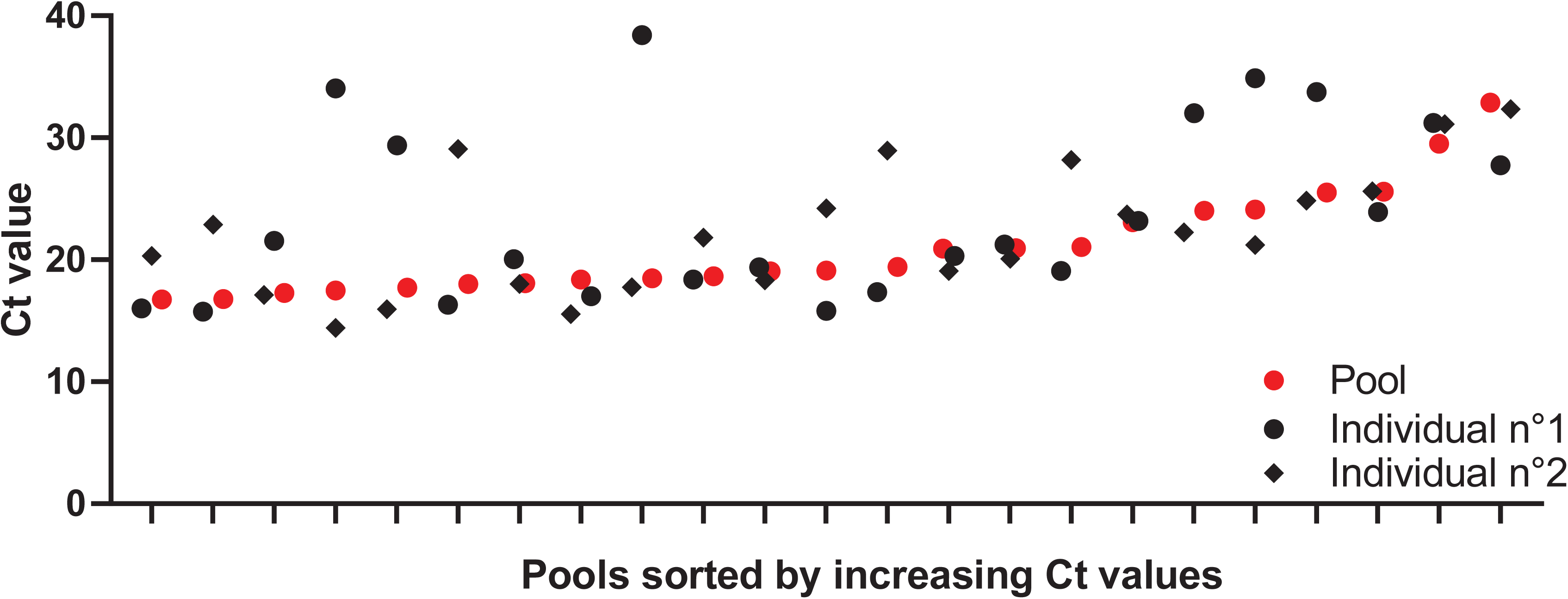
Graph showing Ct values for the detection of SARS-CoV-2 E gene in pools, and of RdRP gene in individual self-collected samples. A) Pools (N=210) containing 1 positive individual sample; B) Pools (N=23) containing 2 positive individual samples; C) Pools (N=5) containing >2 positive individual samples.

RNA extracted from individual samples of the first 50 positive pools reported in French Polynesia (among those containing only one positive individual sample) were also tested for the detection of the SARS-CoV-2 E gene (Figure 4). A mean difference of 1.17 Ct (CI 95% 0.93–1.41) was found between positive individual and pooled samples, following RNA extraction from 200 µL and 50 µL of sample, respectively.

**Fig 4.**
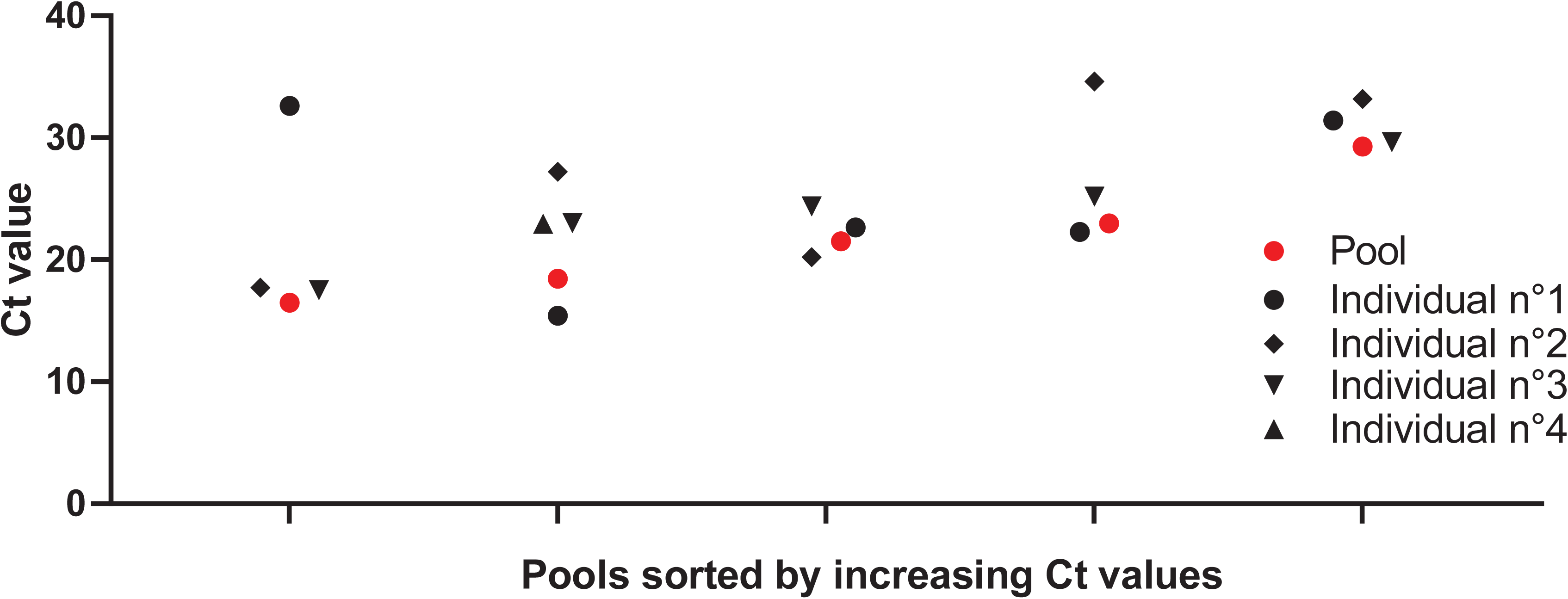
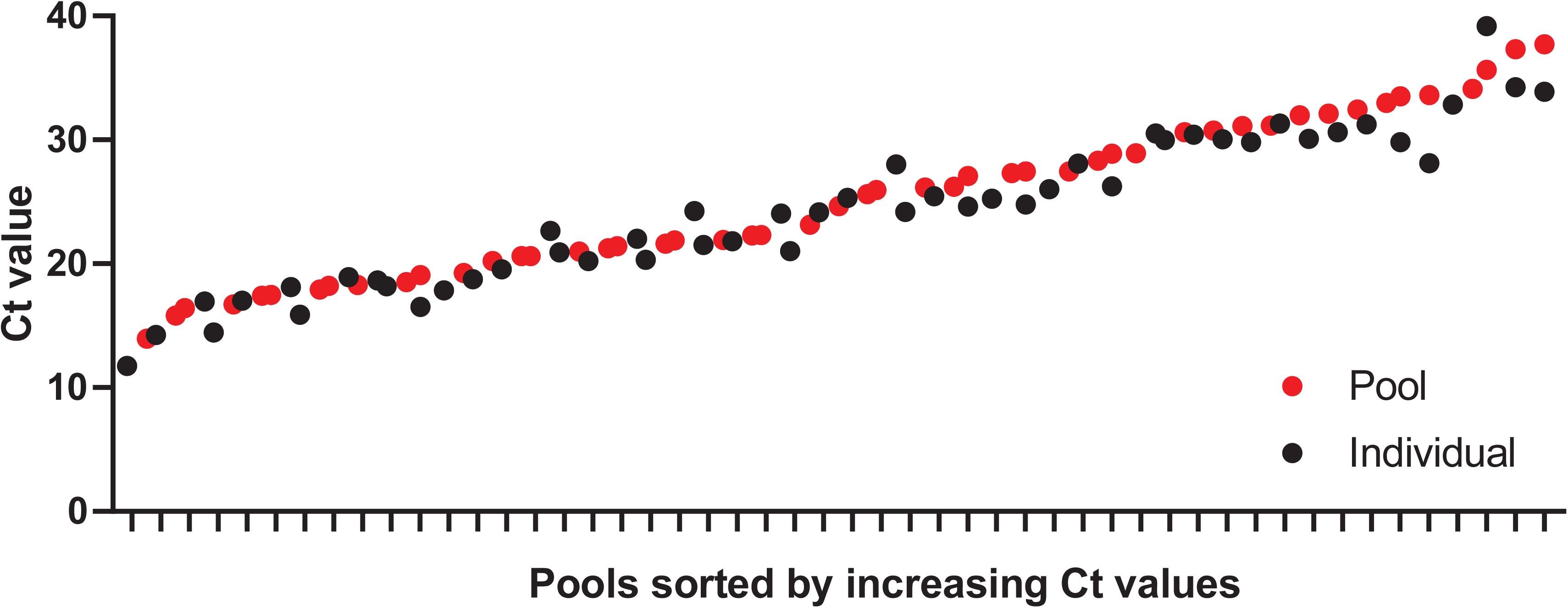
Graph showing Ct values for the detection of SARS-CoV-2 E gene in positive individual and pooled self-collected samples.

### Detection of variants

Among 143 positive individual samples self-collected by travelers from October 20^th^, 2020, 8 were also found positive using variants-specific amplification kit. All travelers had arrived from Metropolitan France. Sequencing of the complete S gene (3,822 bp) from 6 samples identified nucleotide mutations specific to the B.1.1.7 UK variant (Table 2). The 2 remaining travelers tested positive using variants-specific amplification kit were closely related to the 2 other travelers for whom sequencing of the S gene revealed the presence of mutations specific to the B.1.1.7 UK variant.

**Table 2.** Mutations specific to the B.1.1.7 UK variant detected in the S gene of SARS-CoV-2 strains isolated from self-collected samples of 6 travelers.

|  | Traveler 1 | Traveler 2 | Traveler 3 | Traveler 4 | Traveler 5 | Traveler 6 |
| --- | --- | --- | --- | --- | --- | --- |
| Date of reception at ILM (month/year) | 12/2020 | 01/2021 | 01/2021 | 02/2021 | 02/2021 | 02/2021 |
| Amino acid substitutions | N501Y | N501Y | N501Y | N501Y | N501Y | N501Y |
|  | A570D | A570D | A570D | A570D | A570D | A570D |
|  | D614G | D614G | D614G | D614G | D614G | D614G |
|  | P681H | P681H | P681H | P681H | P681H | P681H |
|  | T716I | T716I | T716I | T716I | T716I | T716I |
|  | S982A | S982A | S982A | S982A | S982A | S982A |
|  | D1118H | D1118H | D1118H | D1118H | D1118H | D1118H |
| Amino acid deletions | H69 | H69 | H69 | H69 | H69 | H69 |
|  | V70 | V70 | V70 | V70 | V70 | V70 |
|  | H144 | H144 | H144 | H144 | H144 | H144 |

## Discussion

Since the emergence of SARS-CoV-2 in China in December 2019, the virus has been circulating worldwide [21]. Several countries, including French Polynesia, rapidly closed their borders to prevent the introduction of new COVID-19 cases [5]. In contrast to other countries in the Pacific which have maintained their borders closed, French Polynesia reopened international air traffic from July 15^th^, 2020, while COVID-free at that time [6, 22], in order to revive tourism-related economic activity.

The requirement to have a negative RT-PCR test for SARS-CoV-2 prior to departure to French Polynesia had been in place since April 2020 for residents returning from abroad. They also had to observe a period of isolation after arrival and perform another RT-PCR test before their discharge. Despite these control measures, 2 travelers tested positive at the end of their period of isolation on June 26^th^, 2020 [23], suggesting that they had been infected during the last days preceding their departure from Metropolitan France. These observations supported the idea that in addition to the pre-departure RT-PCR test, a second post-arrival test was needed to detect travelers who may have been infected just before departure.

The COV-CHECK PORINETIA surveillance strategy, consisting in testing the samples self-collected by travelers 4 days after their arrival, enabled to detect SARS-CoV-2 infection in 273 travelers among 59,490 travelers tested between July 15^th^, 2020, and February 15^th^, 2021. Consistent with the results of a previous study showing that up to 11 individual samples could be pooled for a SARS-CoV-2 prevalence of 1 % [19], the prevalence of SARS-CoV-2 infections found at 0.46 % in travelers confirms that our pooling strategy including up to 10 different self-collected samples was relevant for screening travelers entering French Polynesia.

Travelers suspected of being infected by SARS-CoV-2 were immediately contacted by the Surveillance office from the Ministry of Health and placed in isolation. A nasopharyngeal swab was collected by a health worker and tested by RT-PCR to confirm the infection. People who had stayed with the suspected cases were also tested. Positive cases were repatriated to Tahiti as soon as possible in order to benefit from appropriate medical care, especially in the event of the appearance of severe symptoms, and also to protect the population of the islands from SARS-CoV-2 contamination. This strategy enabled the early detection of the first case of COVID-19 since the re-opening of the borders on a cruise ship, and prevented the contamination of other passengers [24].

The COV-CHECK PORINETIA surveillance strategy had some limitations. Indeed, of the 61,397 travelers who were given a self-test kit on arrival, 1,907 (3.11%) did not return their self-collected samples, despite reminders. In addition, since self-collection was not supervised by a health worker, it was impossible to check whether the travelers had correctly collected their samples by strictly following the instructions supplied with the kit. Finally, the pooling samples strategy resulted in a slight loss of sensitivity of the RT-PCR assay compared to individual samples testing (on average 1.17 Ct for the detection of the E gene), probably related to the volume of sample extracted (50 µL versus 200 µL, respectively).

Despite these limitations, the surveillance strategy contributed to protect most remote French Polynesia islands with limited health infrastructures from the introduction of SARS-CoV-2 [25]. In addition to the surveillance strategy implemented since July 15^th^, 2020, travelers were required to apply barrier measures during their stay, thus limiting the risk of contamination of the islands’ population, and to report to the health authorities any symptoms suggestive of COVID-19. Despite these obligations, an asymptomatic traveler whose self-collected sample tested positive but who had not complied with the barrier measures during the 4 days preceding the test generated the first cluster of COVID-19 cases in Tahiti [6]. As of February 15^th^, 2021, 18,293 cases of COVID-19 and 135 deaths related to SARS-CoV-2 infection were recorded [26].

Given the worldwide emergence of several variants of SARS-CoV-2 with an estimated increased transmissibility of up to 70% [27], self-collected samples of travelers that had been found positive from October 20^th^, 2020, using SARS-CoV-2 specific RT-PCR, were retrospectively tested with a variant-specific kit. Subsequent sequencing of the S gene revealed that at least 6 travelers had been infected with the UK variant before entering French Polynesia. Rapid isolation of these travelers prevented the wide spread of the variant, with only 4 secondary cases reported as of February 23^rd^, 2021 [25].

## Conclusion

Depending on the evolution of the epidemiological context of COVID-19 in French Polynesia, the strategy consisting in pooling samples self-collected by travelers was initially used to detect the introduction of SARS-CoV-2 in a COVID-free area with limited medical staff and material resources, then more recently to prevent the emergence of variants. Our results showed that pooling samples had a minor impact on the sensitivity of the RT-PCR test. In order to facilitate SARS-CoV-2 screening in the population, more and more countries have validated the use of self-collected samples [28-30]. Therefore, self-collection and pooling approaches should be considered to prevent, or at least limit, the introduction of COVID-19 cases when tourist trade resumes in countries whose borders are currently closed, and for mass population screening in countries with active SARS-CoV-2 circulation.

## Data Availability

I confirm that all data referred to in the manuscript are available.

## Acknowledgements

We acknowledge the staff of the Ministry of Health, the Ministry of Tourism, Tahiti-Faa’a airport, and the Institut Louis Malardé for their involvement in the COV-CHECK PORINETIA surveillance system. We also thank the staff of hotels and guest houses for participating in receiving samples and sending them to health care centers. Finally, we are grateful to the Laboratory for Urgent Response to Biological Threats at the Institut Pasteur of Paris for providing primers for the sequencing of the SARS-CoV-2 genome. We especially thank Marie Solignac (Institut Louis Malardé) for designing the map of French Polynesia.

## Author contributions

MA, IT, AT, VR and VMCL designed the surveillance protocol. MA, IT and VMCL implemented the surveillance protocol. MA and IT were responsible for the distribution, collection and monitoring of the self-sampling kits. VR and TMH participated in the implementation of the surveillance protocol and sample processing. KC participated in the monitoring of self-collected samples and data collection. AT and VR designed the pooling protocol. AT supervised the experimentations to validate the protocols of pooling, RNA extraction, RT-PCR and sequencing. AT, FD and TP performed pooling and molecular testing of self-collected samples. TP sequenced the SARS-CoV-2 genome and AT analyzed the data. MA wrote the first version of the manuscript. MA and IT performed data analysis. All authors critically reviewed the manusript.

## Supporting information

**S1 File: Documents provided with the self-sampling kit**.

**S1 Table: RT-PCR results (Ct values) for the detection of SARS-CoV-2 E and RdRP genes in positive individual and pooled samples self-collected by travelers from July 15**^**th**^, **2020, to February 15**^**th**^, **2021**.

## COVID-19: INFORMATION FOR TRAVELERS ENTERING FRENCH POLYNESIA

Dear traveler,

Welcome to French Polynesia.

**In order to protect our country from the introduction of coronavirus SARS-CoV-2, a mandatory health surveillance system for the travelers has been implemented by the country (Arrêté n° 525 CM of May 13, 2020)**.

Before your departure, you were asked to provide a negative test for SARS-CoV-2 performed during the 3 days preceding the flight, and to fill out a digital health commitment form on the website www.etis.pf. The information collected on this form should enable to retrace your itinerary if you develop the disease during your stay in French Polynesia.

As the test performed before your departure does not completely exclude the risk that you are infected by SARS-CoV-2, **another test (free of charge) will be carried out during the week following your arrival in French Polynesia, for all travelers aged at least 6 years old**.

This test consists in an oral and nasal swab performed by yourself, at the date* indicated on the envelope containing the kit that you are issued upon arrival in French Polynesia (see self-sampling instructions).

➢ At the date of the test, if you are staying in any accommodation or cruise ship participating in the surveillance of travelers, as soon as the sample collection is performed, you drop it at the reception of the establishment that will be responsible for the delivery to a health care center as listed in the appendix.
➢ Otherwise, you will quickly drop your sample at a health care center or directly at the Institut Louis Malardé in Papeete, at opening hours indicated in the appendix.

*\*If you are staying on an island served less than 3 times a week by plane, please contact the island’s health care center to adjust the collection date according to the next flight*.

This test being carried out as part of a surveillance system, the result will not be communicated to you. However, if a SARS-CoV-2 infection is suspected, you will be contacted by the Health surveillance office of the Direction de la santé.

**Reminders**

If during your stay you have at least one of the symptoms of Covid-19 (fever, cough, sore throat, headache, diarrhea, breathing difficulties, body aches, loss of taste or smell), contact immediately the **reporting platform** at the following number: **(+689) 40 455 000**. If you have sign s of severity, call directly the **SAMU at (+689) 15**.

Throughout your stay in French Polynesia, apply scrupulously the **barrier measures**: respect for physical distance of at least 1 meter, wearing a mask in public areas, and regular cleaning of the hands with soap or hand disinfection with a hydro-alcoholic solution.

## COVID-19 : SURVEILLANCE OF TRAVELERS IN FRENCH POLYNESIA SELF-SAMPLING INSTRUCTIONS

**Sampling kit components**

« Biohazard » bag

Absorbant (leave into the bag)

Swab for oral sampling

Swab for nasal sampling

Tube with transport liquid inside

« Barcode » bag

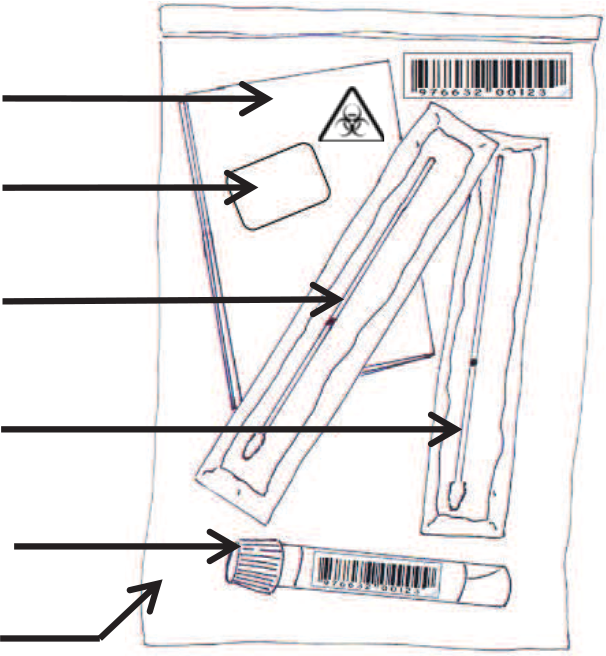

**Instructions for the nasal sampling**

**Make sure not to eat, drink, smoke or brush your teeth at least 20 minutes before the sampling**.

1. Blow your nose with a clean tissue.
2. Wash your hands (with soap or hydro-alcoholic solution).
3. Open the tube containing the transport liquid.
4. Place this tube in a small, stable container (glass, cup) to avoid spilling it.
5. Remove the swab for the nasal sampling from the wrapper by holding the plastic handle. **Do not touch the cotton tip!**
6. Insert the entire cotton tip of the swab into one nostril until you feel a bit of resistance, then rub it in a circle around your nostril 4 times. Repeat the previous action in the second nostril using the same swab.
7. Break the swab at the break point. **Do not touch the cotton tip!**
8. Put the cotton end of the swab in the tube containing the transport liquid (the cotton end must be immersed in the liquid).

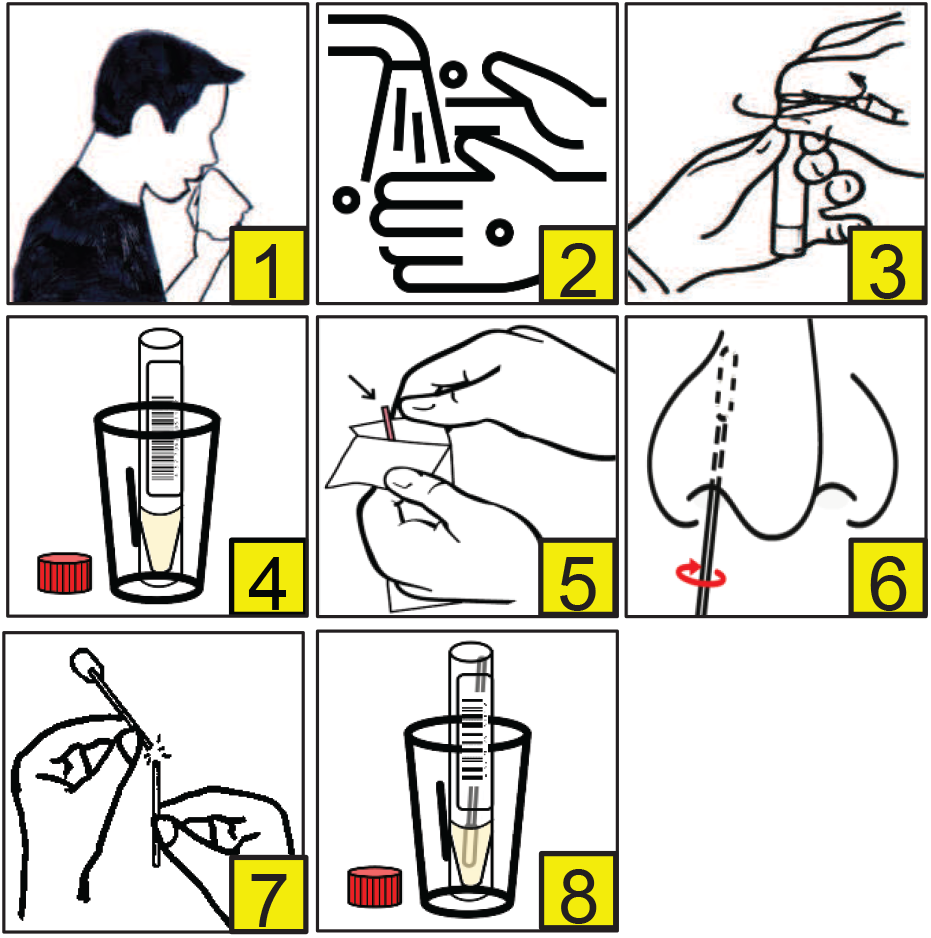

**Instructions for the oral sampling**

9. Take a deep breath then cough 5 times in your elbow.
10. Remove the swab for the oral sampling from the wrapper by holding the plastic handle. **Do not touch the cotton tip!**
11. Insert the cotton tip of the swab into the mouth. Rub several times inside of both cheeks, the top and the bottom of the tongue, the palate, and finally the lower and upper gums (make sure the cotton is well soaked in saliva).
12. Break the swab at the break point. **Do not touch the cotton tip!**
13. Put the cotton end of the swab **in the tube containing the transport liquid previously used for the nasal sampling**.
14. Close the tube by firmly screwing the cap.
15. Turn the tube 3 times to mix the samples impregnated on the cotton with the liquid.
16. Put the tube in the « biohazard » bag then make sure to close the bag tightly (do not remove the absorbant).
17. Put the « biohazard » bag in the « barcode » bag and seal it (do net remove the barcodes stuck on the bag). Store the bag in the refrigerator (between 4°C and 8°C).
18. Give the bag to the referent of your accommodation or drop it in a health care center (see the list provided in the appendix).

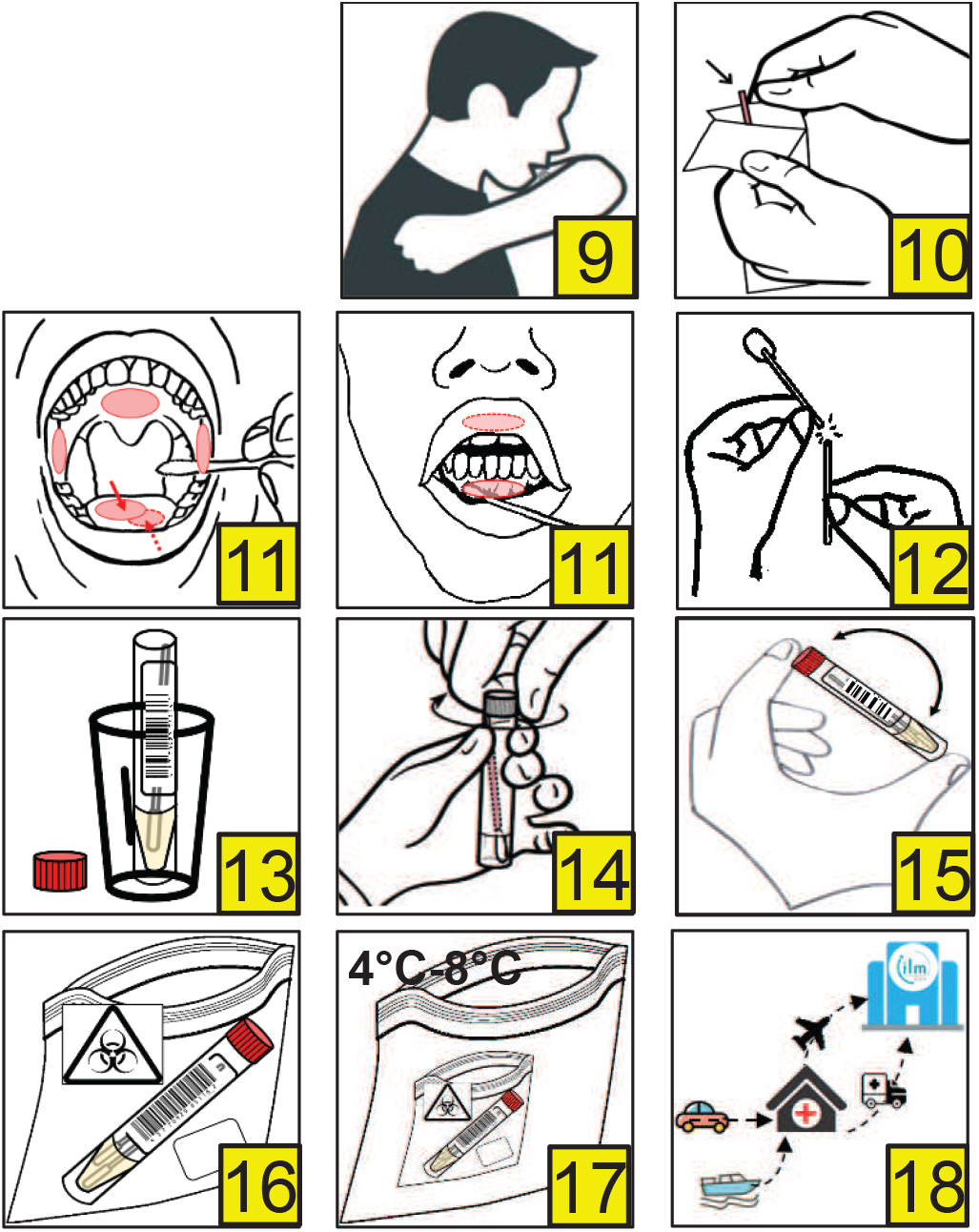

List of health care centers :

➢ **Tahiti**
  - Institut Louis Malardé, Papeete – Tel: (+689) 40 416 459. Open from Monday to Friday from 6.30am to 4pm and Saturday from 8am to 12pm
  - Dispensaire de Papeete – Tel: (+689) 40 549 838. Open from Monday to Thursday 7am to 3pm, Friday from 7am to 2pm
  - Dispensaire de Faa’a – Tel: (+689) 40 850 857. Open from Monday to Thursday 7am to 3pm, Friday from 7am to 2pm
  - Dispensaire de Punaauia – Tel (+689) 40 503 770. Open from Monday to Thursday 7am to 3pm, Friday from 7am to 2pm
  - Dispensaire de Paea – Tel: (+689) 40 533 259. Open from Monday to Thursday 7am to 3pm, Friday from 7am to 2pm
  - Dispensaire de Papara – Tel: (+689) 40 574 787. Open from Monday to Thursday 7am to 3pm, Friday from 7am to 2pm
  - Dispensaire de Teva I Uta – Tel: (+689) 40 547 270. Open from Monday to Thursday 7am to 3pm, Friday from 7am to 2pm
  - Dispensaire de Arue – Tel: (+689) 40 455 959. Open from Monday to Thursday 7am to 3pm, Friday from 7am to 2pm
  - Dispensaire de Mahina – Tel: (+689) 40 481 885. Open from Monday to Thursday 7am to 3pm, Friday from 7am to 2pm
  - Dispensaire de Papenoo – Tel: (+689) 40 423 671. Open from Monday to Thursday 7am to 3pm, Friday from 7am to 2pm
  - Dispensaire de Tiarei – Tel: (+689) 40 521 633. Open from Monday to Thursday 7am to 3pm, Friday from 7am to 2pm
  - Dispensaire de Hiti’aa – Tel: (+689) 40 521 019. Open from Monday to Thursday 7am to 3pm, Friday from 7am to 2pm
  - Hôpital de Taravao – Tel: (+689) 40 547 782. Open from Monday to Thursday 7:30am to 3:30pm, Friday from 7:30am to 2:30pm
➢ **Moorea**
  - Hôpital d’Afareaitu – Tel: (+689) 40 552 222. Open from Monday to Thursday 7am to 3pm, Friday from 7am to 2pm
➢ **Raiatea**
  - Hôpital d’Uturoa – Tel: (+689) 40 600 800. Open from Monday to Thursday 7:30am to 3:30pm, Friday from 7:30am to 2:30pm
➢ **Tahaa**
  - Centre médical – Tel: (+689) 40 656 331. Open from Monday to Thursday 7:30am to 3:30pm, Friday from 7:30am to 2:30pm
  - Centre médical – Tel: (+689) 40 656 751. Open on Tuesday from 7:30am to 12:00am
➢ **Huahine**
  - Dispensaire – Tel: (+689) 40 688 248. Open from Monday to Thursday 7:30am to 3:30pm, Friday from 7:30am to 2:30pm
➢ **Bora Bora**
  - Dispensaire – Tel: (+689) 40 677 077. Open from Monday to Thursday 7:30am to 3:30pm, Friday from 7:30am to 2:30pm
➢ **Maupiti**
  - Infirmerie – Tel: (+689) 40 678 018. Open from Monday to Thursday 7:30am to 3:30pm, Friday from 7:30am to 2:30pm
➢ **Nuku Hiva**
  - Hôpital Taiohae – Tel: (+689) 40 910 200. Open from Monday to Thursday 7:30am to 3:30pm, Friday from 7:30am to 2:30pm
  - Infirmerie de Taipivai – Tel: (+689) 40 920 142. Open from Monday to Friday 7:30am to 1:30pm
  - Infirmerie de Hatiheu – Tel: (+689) 40 920 143. Open from Monday to Friday 7:30am to 1:30pm
➢ **Ua Huka**
  - Infirmerie de Hane – Tel: (+689) 40 926 058. Open from Monday to Friday 7:30am to 1:30pm
➢ **Ua Pou**
  - Centre médical de Hakahau – Tel: (+689) 40 925 375. Open from Monday to Thursday 7:30am to 3:30pm, Friday from 7:30am to 2:30pm
  - Infirmerie de Hakamaii – Tel: (+689) 40 925 299. Open from Monday to Friday 7:30am to 1:30pm
  - Infirmerie de Hakatao – Tel: (+689) 40 925 104. Open from Monday to Friday 7:30am to 1:30pm
➢ **Hiva Oa**
  - Centre médical de Atuona – Tel: (+689) 40 927 375. Open from Monday to Thursday 7:30am to 3:30pm, Friday from 7:30am to 2:30pm
  - Infirmerie de Paumau – Tel: (+689) 40 927 496. Open from Monday to Friday 7:30am to 1:30pm
➢ **Tahuata**
  - Infirmerie de Vaitahu – Tel : (+689) 40 929 227. Open from Monday to Friday 7:30am to 1:30pm
➢ **Fatu Hiva**
  - Infirmerie de Omoa – Tel: (+689) 40 928 036. Open from Monday to Friday 7:30am to 1:30pm
➢ **Tubuai**
  - Centre médical de Mataura – Tel: (+689) 40 932 250. Open from Monday to Thursday 7:30am to 3:30pm, Friday from 7:30am to 2:30pm
➢ **Rurutu**
  - Centre médical de Moerai – Tel: (+689) 40 930 440. Open from Monday to Thursday 7:30am to 3:30pm, Friday from 7:30am to 2:30pm
➢ **Rimatara**
  - Infirmerie de Amaru – Tel: (+689) 40 944 270. Open from Monday to Thursday 7:30am to 3:30pm, Friday from 7:30am to 2:30pm
➢ **Raivavae**
  - Infirmerie de Rairua – Tel: (+689) 40 95 42 31. Open from Monday to Thursday 7:30am to 3:30pm, Friday from 7:30am to 2:30pm
➢ **Rangiroa**
  - Centre médical de Avatoru – Tel: (+689) 40 960 325.
  - Infirmerie de Tiputa – Tel: (+689) 40 967 396. Open from Monday to Friday 7:30am to 12:00am. Emergency 24h/24, everyday
➢ **Hao**
  - Centre médical – Tel: (+689) 40 970 513.
➢ **Makemo**
  - Centre médical – Tel: (+689) 40 980 325. Open from Monday to Thursday 7:30am to 3:30pm, Friday from 7:30am to 2:30pm
  - Infirmerie – Tel: (+689) 40 980 325. Open from Monday to Friday 7:30am to 12:00am. Emergency 24h/24, everyday
➢ **Gambier**
  - Centre médical de Rikitea – Tel: (+689) 40 978 216. Open from Monday to Thursday 7:30am to 3:30pm, Friday from 7:30am to 2:30pm
➢ **Ahe**
  - Infirmerie – Tel: (+689) 40 964 403. Open from Monday to Friday 7:30am to 12:00am. Emergency 24h/24, everyday
➢ **Anaa**
  - Infirmerie – Tel: (+689) 40 983 204. Open from Monday to Friday 7:30am to 12:00am. Emergency 24h/24, everyday
➢ **Arutua**
  - Infirmerie – Tel: (+689) 40 965 300. Open from Monday to Friday 7:30am to 12:00am. Emergency 24h/24, everyday
➢ **Fakarava**
  - Infirmerie – Tel: (+689) 40 984 224. Open from Monday to Friday 7:30am to 12:00am. Emergency 24h/24, everyday
➢ **Manihi**
  - Infirmerie – Tel: (689) 40 964 136. Open from Monday to Friday 7:30am to 12:00am. Emergency 24h/24, everyday
➢ **Napuka**
  - Infirmerie – Tel : (+689) 40 973 260. Open from Monday to Friday 7:30am to 12:00am. Emergency 24h/24, everyday
➢ **Reao**
  - Infirmerie – Tel: (+689) 40 969 041. Open from Monday to Friday 7:30am to 12:00am. Emergency 24h/24, everyday
➢ **Takapoto**
  - Infirmerie – Tel: (+689) 40 986 486. Open from Monday to Friday 7:30am to 12:00am. Emergency 24h/24, everyday
➢ **Takaroa**
  - Infirmerie – Tel: (+689) 40 982 263. Open from Monday to Friday 7:30am to 12:00am. Emergency 24h/24, everyday
➢ **Tikehau**
  - Infirmerie – Tel: (+689) 40 962 349. Open from Monday to Friday 7:30am to 12:00am. Emergency 24h/24, everyday

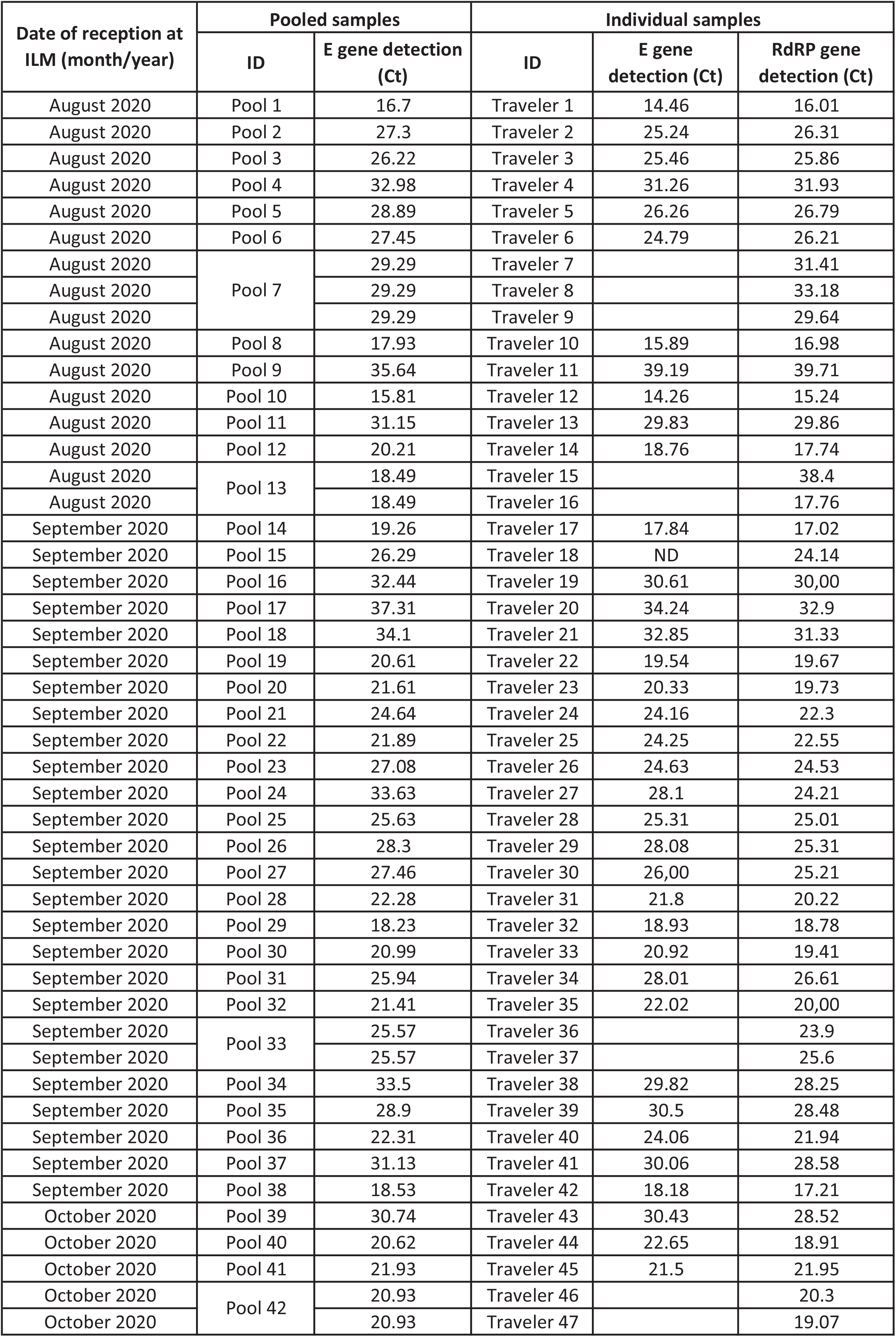
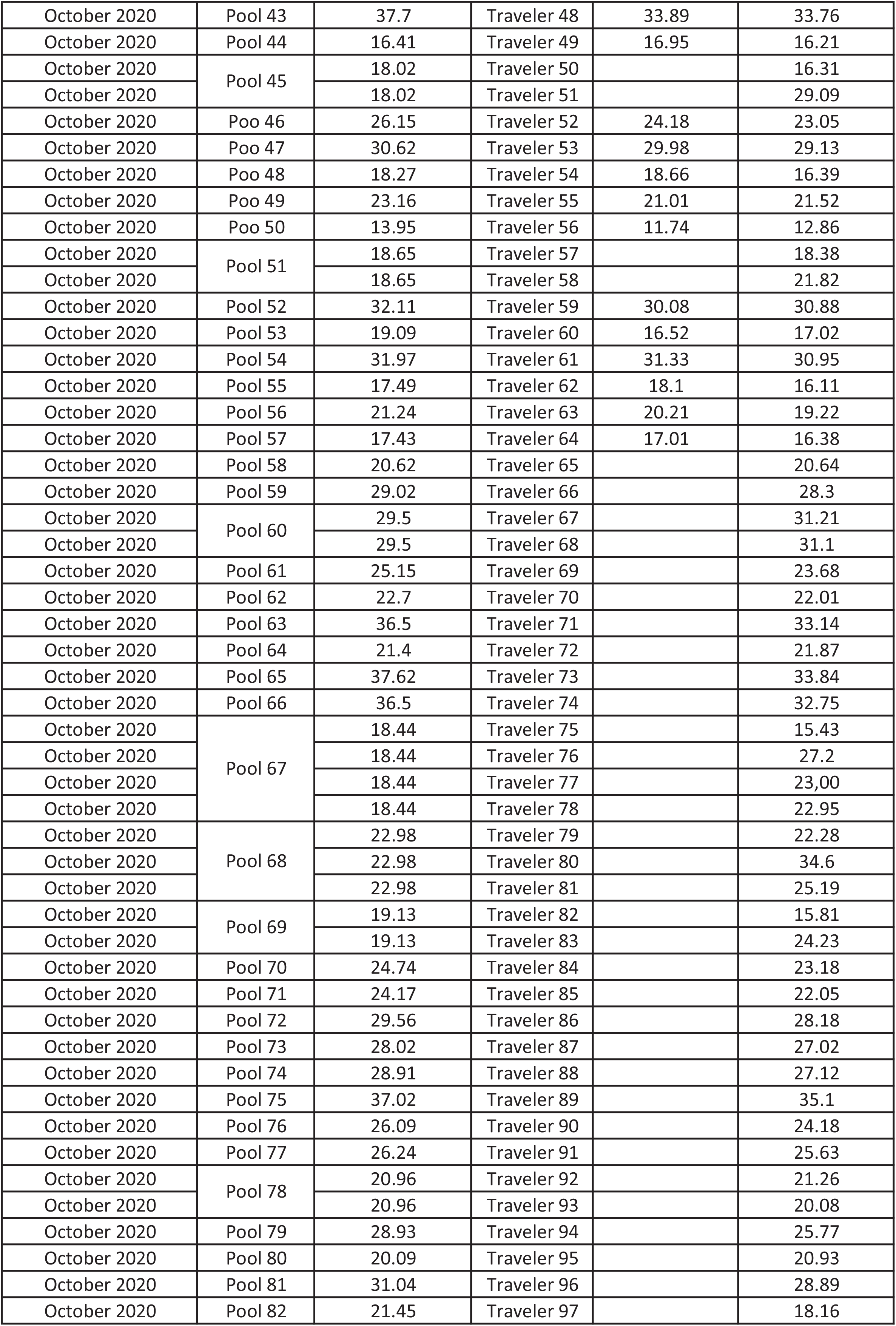
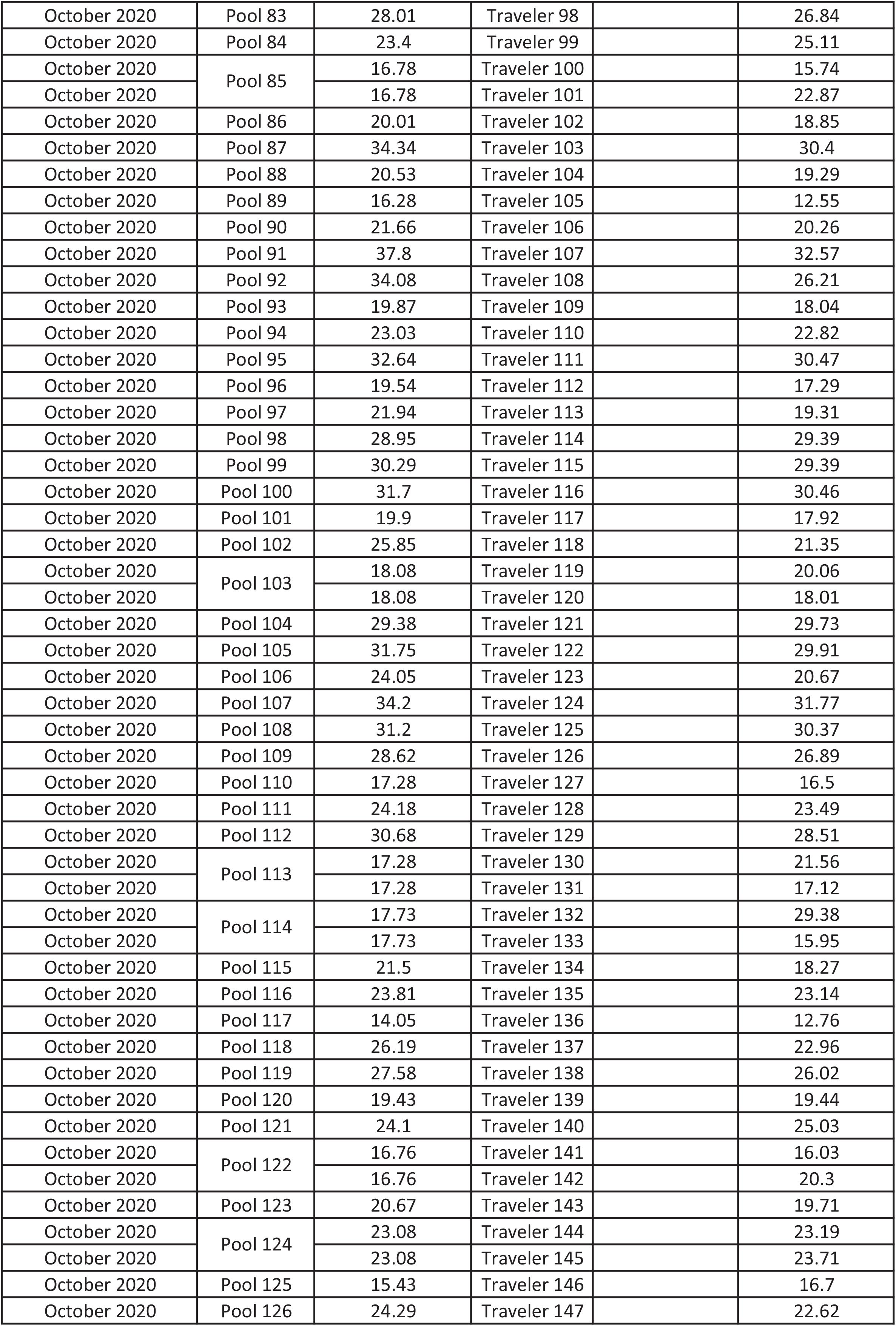
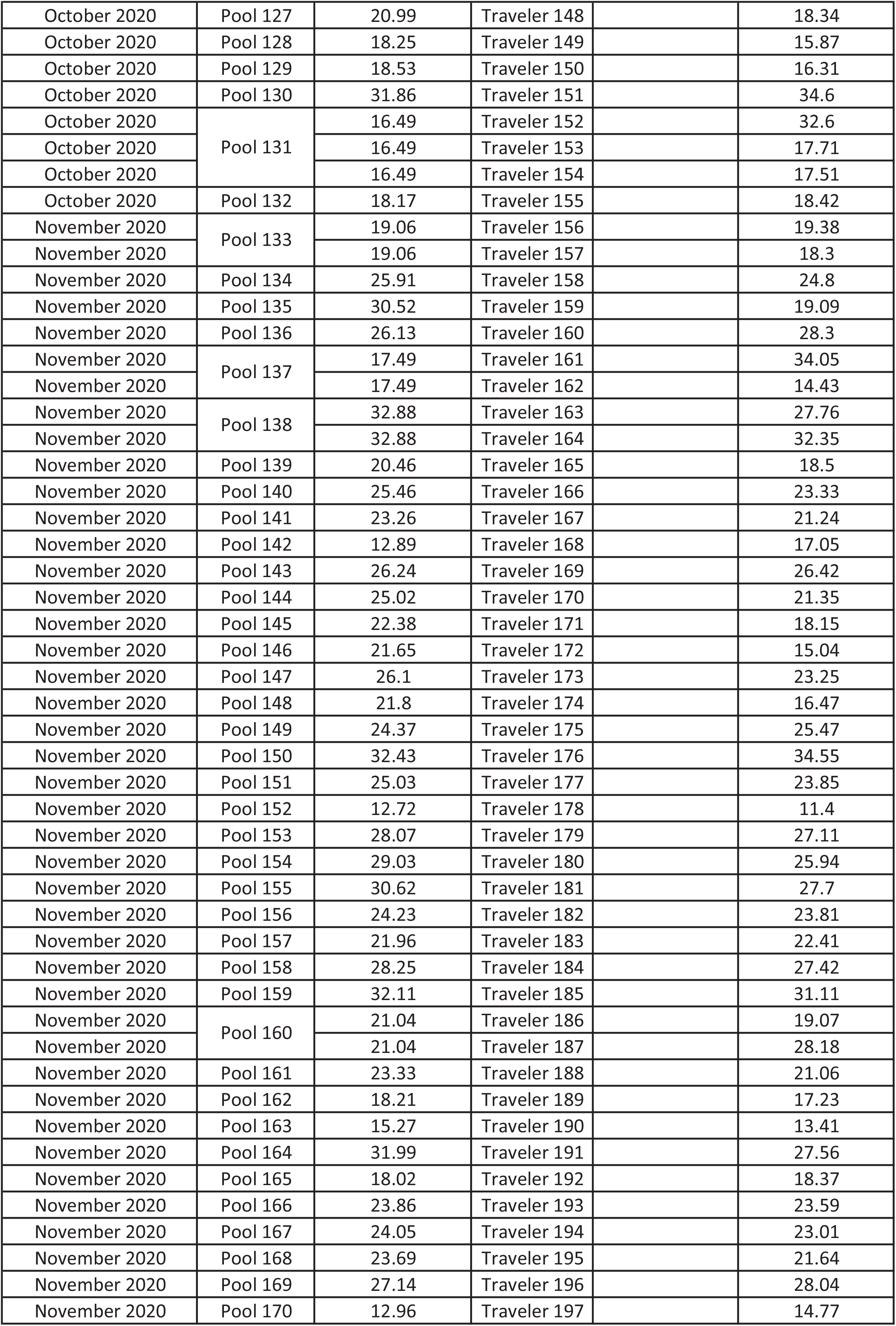
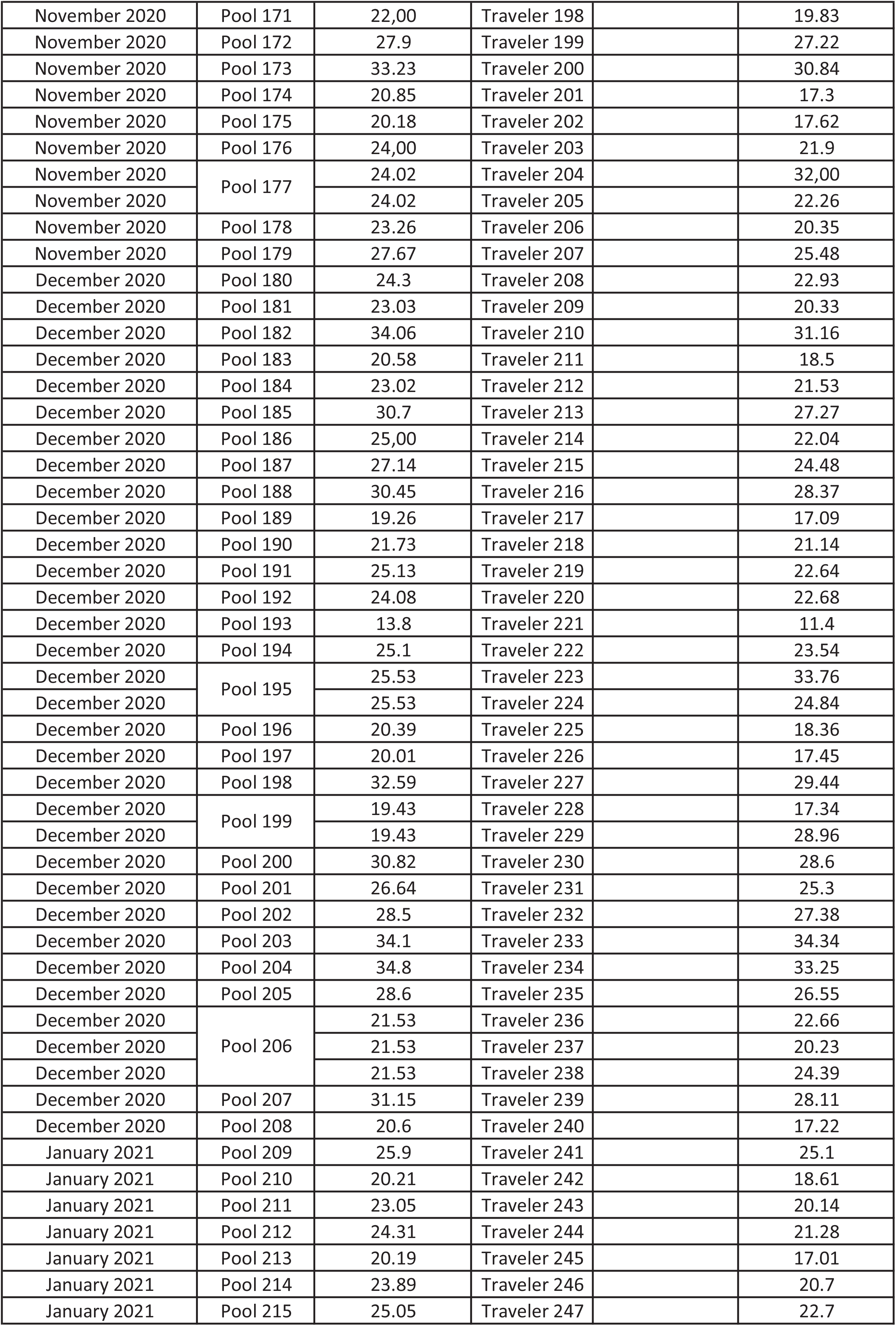
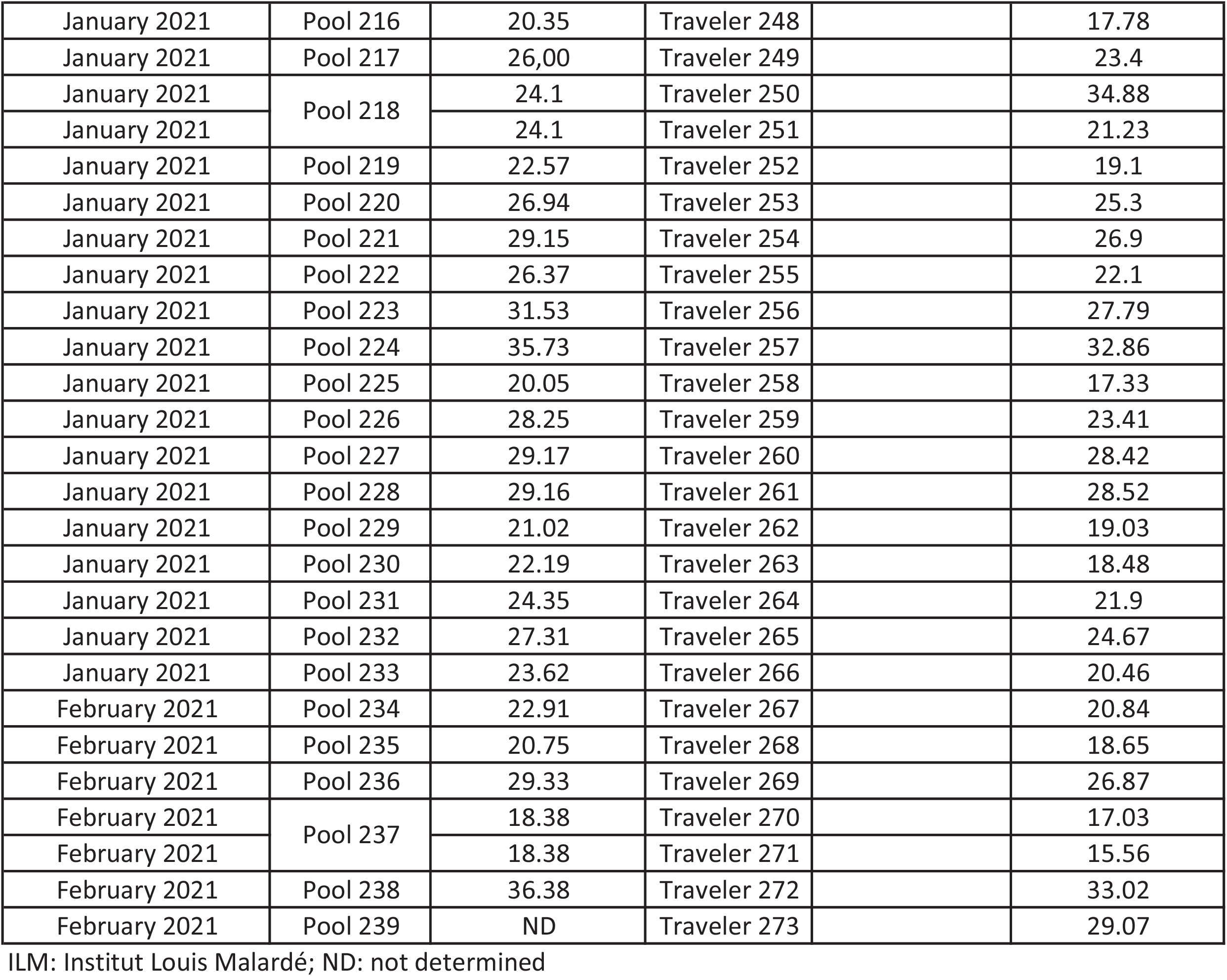

## Notes

### Competing Interest Statement

The authors have declared no competing interest.

### Funding Statement

The COV-CHECK PORINETIA surveillance strategy has been funded by the Government of French Polynesia.

### Author Declarations

The approval for the use of data produced within the framework of the COV-CHECK PORINETIA surveillance strategy was obtained from the ethics committee of French Polynesia (No. 90 CEPF of June 15th, 2021)

